# Characteristics of Patients Referred to a Cardiovascular Disease Clinic for Post-Acute Sequelae of SARS-CoV-2 Infection

**DOI:** 10.1101/2021.12.04.21267294

**Authors:** Stephen Y. Wang, Philip Adejumo, Claudia See, Oyere K. Onuma, Edward J. Miller, Erica S. Spatz

## Abstract

There is limited literature on the cardiovascular manifestations of post-acute sequelae of SARS-CoV-2 infection (PASC). We aimed to describe the characteristics, diagnostic evaluations, and cardiac diagnoses in patients referred to a cardiovascular disease clinic designed for patients with PASC from May 2020 to September 2021. Of 126 patients, average age was 46 years (range 19-81 years), 43 (34%) were male. Patients presented on average five months after COVID-19 diagnosis. 30 (24%) patients were hospitalized for acute COVID-19. Severity of acute COVID-19 was mild in 37%, moderate in 41%, severe in 11%, and critical in 9%. Patients were also followed for PASC by pulmonology (53%), neurology (33%), otolaryngology (11%), and rheumatology (7%). Forty-three patients (34%) did not have significant comorbidities. The most common symptoms were dyspnea (52%), chest pain/pressure (48%), palpitations (44%), and fatigue (42%), commonly associated with exertion or exercise intolerance. The following cardiovascular diagnoses were identified: nonischemic cardiomyopathy (5%); new ischemia (3%); coronary vasospasm (2%); new atrial fibrillation (2%), new supraventricular tachycardia (2%); myocardial involvement (15%) by cardiac MRI, characterized by late gadolinium enhancement (LGE; 60%) or inflammation (48%). The remaining 97 patients (77%) exhibited common symptoms of fatigue, dyspnea on exertion, tachycardia, or chest pain, which we termed “cardiovascular PASC syndrome.” Three of these people met criteria for postural orthostatic tachycardia syndrome. Lower severity of acute COVID-19 was a significant predictor of cardiovascular PASC syndrome. In this cohort of patients referred to cardiology for PASC, 23% had a new diagnosis, but most displayed a pattern of symptoms associated with exercise intolerance.

## INTRODUCTION

Post-acute sequelae of SARS-CoV-2 infection (PASC), also known as Long COVID, is a long-term manifestation of acute coronavirus disease 2019 (COVID-19), affecting up to ten to thirty percent of patients, and defined as emergent or persistent symptoms extending beyond four weeks from onset of acute illness.^1^ Despite growing reports of cardiopulmonary symptoms associated with PASC, the literature on cardiovascular manifestations in this post-acute period remains limited.

## METHODS

This was an observational study of all consecutive patients referred to the Yale cardiovascular clinic for PASC (part of the Comp**RE**hensive Post-**COV**ID Cent**ER** at **Y**ale,^2^ RECOVERY) between May 2020 and September 2021. Clinical data were manually extracted from the electronic medical record. Diagnostic evaluation included baseline vital signs, cardiopulmonary physical examination, ECG, laboratory studies, and echocardiogram. Initially, all patients had a NT-proBNP, troponin, d-dimer, and CRP – however, these tests were overwhelmingly normal and subsequently only checked based on clinical suspicion. An active stand test for postural orthostatic tachycardia syndrome (POTS) was performed in patients with orthostatic intolerance (blood pressure and heart rate measured after 5 minutes in the supine position and with 10 minutes of standing). Further evaluation was guided by clinical judgement to: a) identify cardiovascular disease pathology, and b) evaluate cardiovascular symptoms not readily attributable to a cardiovascular diagnosis, but which may have a cardiopulmonary etiology. We describe the clinical characteristics and diagnostic yield of testing, along with predictors of having cardiovascular symptoms without an identified diagnosis. The study was approved by the Human Investigation Committee of Yale University.

## RESULTS

Of 126 patients, average age was 44 years (range 19-74 years), 9% were ≥65 years old, and 83 (66%) were female. Thirty patients (24%) had been hospitalized for acute COVID-19 (**Table**). Severity of acute COVID-19 was mild in 37%, moderate in 41%, severe in 11%, and critical in 9%.^3^ Patients presented on average (SD) 5.1 months (3.8 months) after acute infection. All but 6 patients had confirmed antigen studies for COVID-19. The most common referrals were from primary care (54%), pulmonology (26%), and cardiology (12%). Patients were also followed for PASC by other specialties: pulmonology (53%), neurology (33%), otolaryngology (11%), and rheumatology (7%). Comorbidities included hypertension (23%), hyperlipidemia (25%), diabetes (17%), and obesity (37%). Twelve patients (10%) had underlying cardiovascular disease, and 40 patients (32%) had underlying pulmonary or rheumatologic disease.

**Table.**
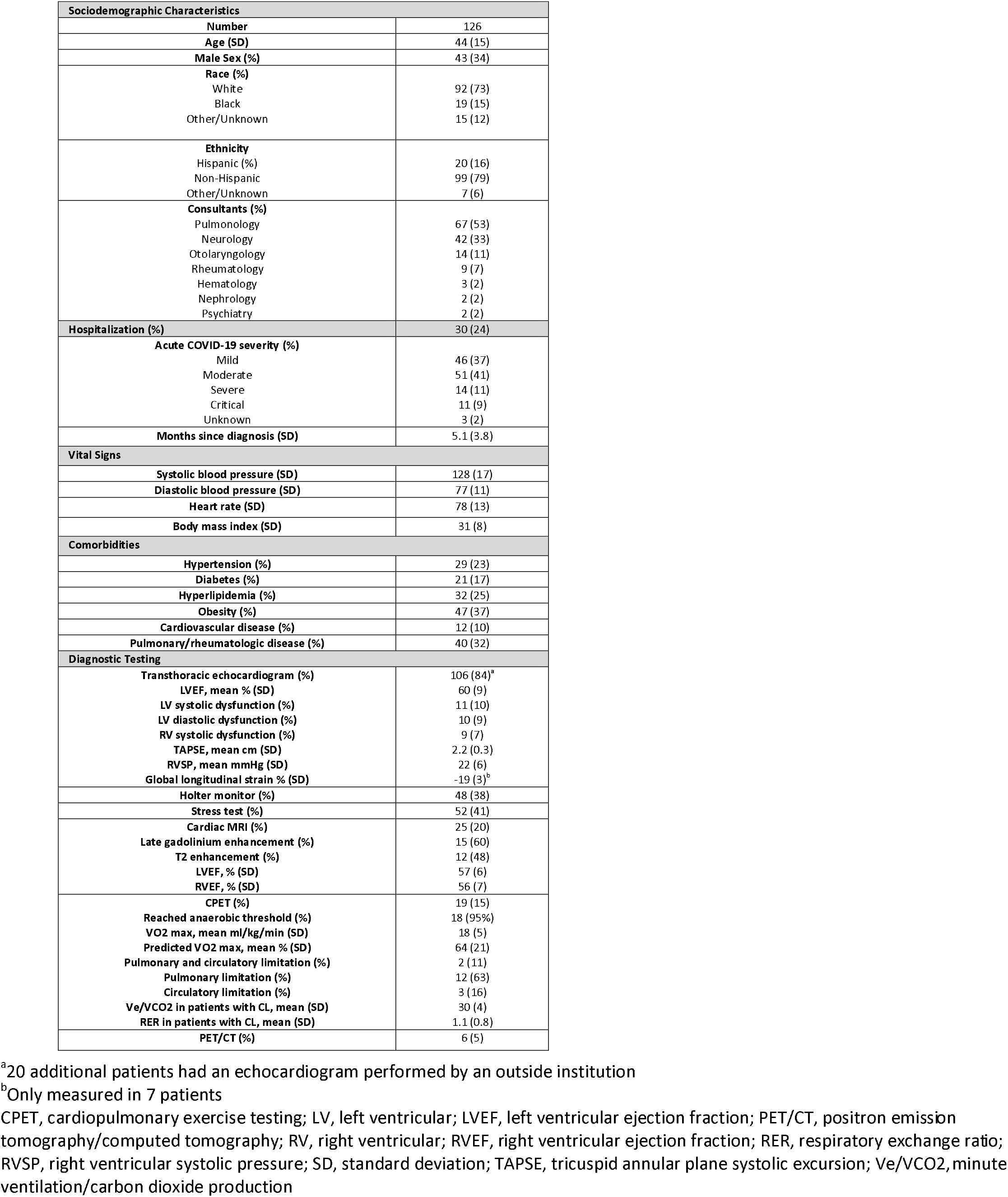
Characteristics of patients at time of presentation to a cardiovascular disease clinic for post-acute sequelae of SARS CoV-2 Infection.

The most common cardiovascular symptoms on presentation included dyspnea (52%, almost always exertional), chest pain/pressure (48%) and palpitations (44%); 84% of patients demonstrated at least one of these cardiopulmonary symptoms (**Figure 1**).

**Figure 1.**
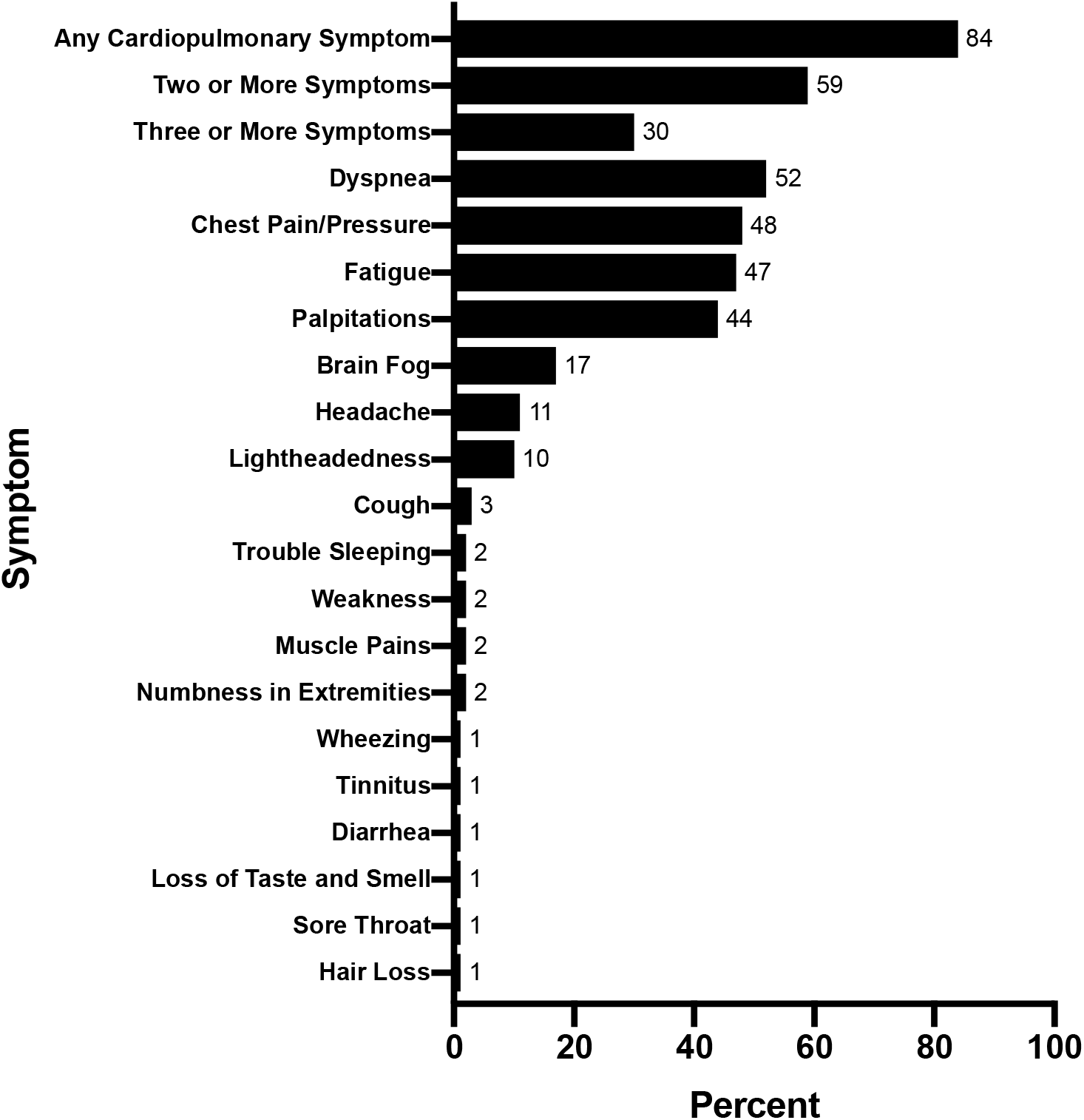
Baseline Symptoms of Patients Referred for Cardiovascular Symptoms associated with Post-Acute Sequelae of SARS-CoV-2 (PASC)

### Diagnostic Evaluation for Cardiovascular Pathology

Echocardiography was performed by the Yale Heart and Vascular Center (n=106); 20 were performed at an outside institution and reported as “normal” (**Table**). Of the remaining 106 patients, the mean left ventricular ejection fraction was 60%. Eleven patients (9%) had evidence of left ventricular systolic dysfunction (no baseline TTEs for comparison), 10 patients (8%) had left ventricular diastolic dysfunction, and 9 patients (7%) had right ventricular systolic dysfunction. Mean right ventricular systolic pressure was 22mmHg, mean TAPSE was 2.2cm, and mean global longitudinal strain (measured in 7 patients) was −19.4%. Forty-eight patients (38%) had a Holter monitor, of whom 43 had non-exertional sinus tachycardia (with symptoms), three had new atrial fibrillation, and two had another supraventricular tachycardia. Four out of 53 patients who underwent stress testing (with SPECT or echocardiogram) had evidence of new ischemia (**Figure 2**). Twenty-five patients (20%) had a cardiac MRI, of whom 76% had late gadolinium enhancement and/or evidence of T2 inflammation, which we diagnosed as myocardial involvement but not acute myocarditis since the presentation was not acute, troponin was negative, and there were no electrocardiographic changes. Other new diagnoses included: nonischemic cardiomyopathy (n=6; 5%) and coronary vasospasm (n=2; 2%).

**Figure 2.**
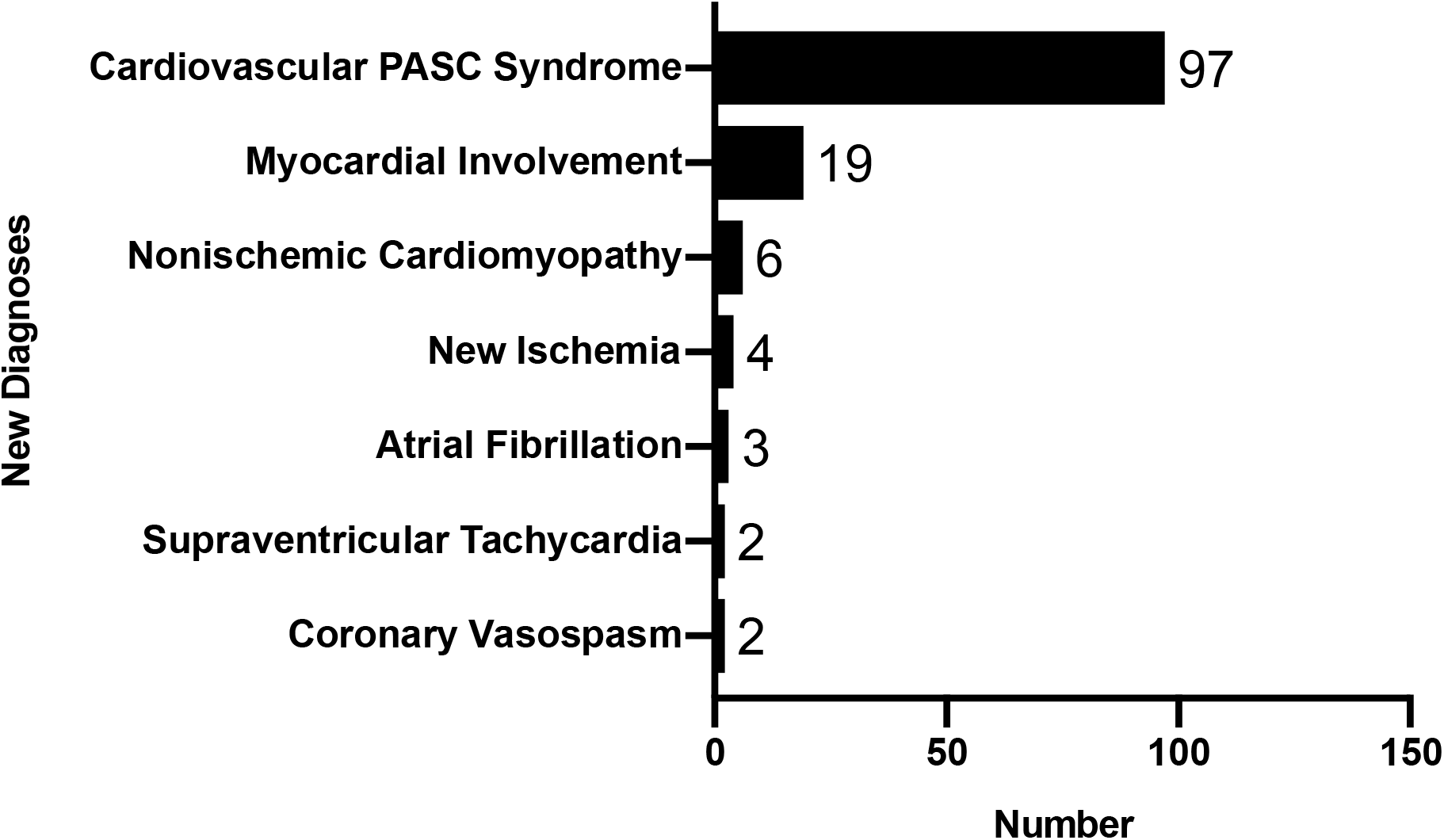
New Diagnoses Identified at Evaluation for Cardiovascular Symptoms associated with Post-Acute Sequelae of SARS-CoV-2 (PASC)^a^ ^a^Numbers do not add up to 126 since some diagnoses overlap in the same patients

### Other Diagnoses and Symptom Patterns

The remaining 97 patients (77%) without new cardiovascular pathology had a “cardiovascular PASC syndrome” of palpitations, chest pain/pressure, fatigue or dyspnea on exertion that limited regular activities. Three patients met criteria for POTS; 1 of whom had a confirmatory tilt table test. Nineteen (15%) underwent cardiopulmonary exercise testing (CPET) to further evaluate symptoms (**Table**). 95% reached anaerobic threshold with a mean max VO2 of 18ml/kg/min. Of those with cardiac limitation, the mean Ve/VCO2 was 30, and the mean peak respiratory exchange ratio was 1.11. Ten patients did not demonstrate any pulmonary limitation, but had varying degrees of cardiac limitation.

Severe (OR=0.24, 95% CI 0.06 – 0.90) and critical (OR=0.22, 95% CI 0.05 – 0.90) severity of acute COVID-19 was associated with a significantly lower odds of cardiovascular PASC syndrome compared with mild severity. Age, sex, and comorbidities were not significantly associated with cardiovascular PASC syndrome.

## DISCUSSION

In this large, consecutive series of patients referred to the cardiovascular disease clinic for PASC, we identified a range of cardiovascular disease pathology. Most patients, however, exhibited a “cardiovascular PASC syndrome” without an underlying diagnosis.

Our study continues to show that cardiovascular manifestations of PASC are common,^4^ affecting primarily non-elderly individuals.^5^ While patients were referred for cardiopulmonary symptoms suspected to have a cardiac etiology, only 23% of evaluations led to a cardiovascular diagnosis, including new cardiomyopathy, ischemia, arrhythmias, and myocardial involvement. Many patients, including those with cardiovascular diagnoses, suffered from exercise intolerance, describing fatigue with activities of daily living and severe post-exertional dyspnea and malaise impairing their ability to work; however, in the majority of cases (77%), a cardiovascular disease was not found. Many of these patients also suffered from “brain fog,” characterized by memory problems, difficulty concentrating, and sleep disturbance. Many also described body aches. These symptoms overlap with myalgic encephalomyelitis-chronic fatigue syndrome (ME-CFS).^6^

COVID-19 can affect the cardiovascular system through several mechanisms, including viral injury, inflammation, dysregulation of ACE-2, along with metabolic and autonomic impairment; however, in a standard clinical evaluation, these mechanisms are challenging to discern.^7^ We did observe downstream physiological dysregulation - including endothelial dysfunction along with cardiometabolic dysfunction – in a subset of patients who underwent invasive coronary vasomotion testing and cardiopulmonary exercise testing. Endothelial dysfunction was directly observed in two patients with ischemia and no obstructive coronary artery disease who developed vasospasm to acetylcholine challenge. For these patients, we prescribed calcium channel blockers, nitrates, statins, and L-arginine. Interestingly, of six patients who underwent stress PET/CT, we detected only one case of microvascular dysfunction. Cardiometabolic impairment was observed on CPET; specifically, more than half of patients had low peak VO2 with normal cardiac and pulmonary function, which may be secondary to bedrest deconditioning and/or impaired oxygen extraction. In bedrest deconditioning, low plasma volume leads to decreased left ventricular filling, cardiac atrophy, and resultant decreased stroke volume which potentiates a compensatory tachycardia to maintain cardiac output.^8^ Fatigue and tachycardia result in less physical activity, which further exacerbates the cycle of deconditioning. Impaired oxygen extraction in the peripheral microcirculation was previously demonstrated on invasive CPET in patients with ME-CFS and more recently with PASC.^9 10^ For all patients with exercise intolerance, we employed principles from the Levine protocol, developed for patients with POTS, which includes structured recumbent exercise with gradual increases in duration, along with hydration, salt loading, compression socks, and elevating the head of the bed.^11^

This study should be interpreted in the context of the following limitations. First, referrals and diagnostic evaluation were not systematic, but may be generalizable to other cardiology practices seeing high volumes of patients with PASC. Second, our understanding of PASC evolved with time, and we eventually reduced the number of diagnostic tests consistent with our growing experience that the overwhelming majority of patients with mild acute COVID-19 infection presenting with exertional dyspnea and tachycardia had no cardiovascular disease pathology; still, all patients had a thorough cardiovascular physical exam, ECG, and echocardiogram. Third, the clinical relevance of myocardial involvement based on cardiac MRI is uncertain. Finally, new diagnoses that better define the PASC cardiovascular syndrome may emerge, although diagnostic algorithms have yet to be developed.

In conclusion, this study provides insight into patients with cardiovascular manifestations of PASC, emphasizing the need for multidisciplinary teams and accelerated research to better phenotype and treat a heterogeneous group of patients.

## Data Availability

All data produced in the present study are available upon reasonable request to the authors

## ACKNOWLEDGMENT

Disclosures: Dr Spatz is a co-PI on a multi-center, Centers for Disease Control and Prevention (CDC)-funded study, “Innovative Support for Patients With SARS-COV2 Infections (COVID-19) Registry (INSPIRE).” She receives funding from the Food and Drug Administration (FDA) as part of the Yale University-Mayo Clinic Center of Excellence in Regulatory Science and Innovation (CERSI). She also receives support from the National Institute on Minority Health and Health Disparities (U54MD010711-01) to study precision-based approaches to diagnosing and preventing hypertension, from the National Institute of Biomedical Imaging and Bioengineering (R01 EB028106-01) to study a cuff-less blood pressure device. The other authors have no relevant disclosures.

